# Early clinical prediction of neurodevelopmental outcome in *KCNQ2*-related disorders

**DOI:** 10.64898/2026.08.06.26359418

**Authors:** Elisabeth Van Boxstael, Charissa Millevert, Megan Hairabedian, Johannes Lemke, Steffen Syrbe, Eugenia Roza, Laura Licchetta, Duccio M Cordelli, Trine B Hammer, Magdalena Krygier, Marta Pietruszka, Safa Mete Dagdas, Pinar Gençpinar, Carmen Fons, Dídac Casas-Alba, Laurence Chaton, Gaëtan Lesca, Irene Valenzuala Palafoll, An-Sofie Schoonjans, Anna Jansen, Tejasvi Nirinjan, Annie Ting-Gee Chiu, Ingrid E Scheffer, Christian Boßelmann, Ludovica Montanucci, Tobias Bruenger, KCNQ2 study group, Dennis Lal, Mathieu Milh, Sarah Weckhuysen

## Abstract

**Objective:** In KCNQ2-related disorders (*KCNQ2*-RD), neurodevelopmental outcome remains variable despite established genotype-phenotype correlations. Our aim is to improve counselling, by developing and internally validating models predicting neurodevelopmental outcomes based on early clinical and genetic features, universally available to clinicians.

**Methods:** We conducted a multicentric retrospective cohort study including 277 individuals carrying a (likely) pathogenic variant in the *KCNQ2* gene, with a minimum follow-up age of three years. Mosaic variants were excluded. The cohort was randomly split into training (70%) and validation (30%) sets. Ten expert selected parameters with minimal missing data were used to train random forest models to predict (i) dichotomous outcomes and (ii) three-category outcomes for cognition, language, and gross motor milestones.

**Results:** Models incorporated seven clinical (neonatal hypotonia, EEG characteristics, age at seizure onset, seizure type, and seizure frequency at onset, prematurity, and sex) and three genetic variables (*de novo* status, exon localisation, and position within known *KCNQ2*-developmental and epileptic encephalopathy (DEE) hotspot regions). Dichotomous models showed the highest predictive performance, with accuracies of 0.83 for normal vs. mild-profound intellectual disability (ID), 0.83 for achievement of first words, and 0.86 for achievement of independent walking. Three-category models remained clinically informative: accuracies were 0.79 for normal vs. mild vs. moderate-profound ID, 0.70 for first words ≤16 months vs. >16 months vs. never, and 0.71 for independent walking ≤18 months vs. >18 months vs. never. The strongest predictors for adverse neurodevelopmental outcomes were presence of hypotonia at birth, seizure onset within the first day of life, multiple seizures per day at onset, tonic seizures at onset, a burst-suppression pattern on EEG at onset, the presence of a *de novo* variant, and variant location within exons 6-7.

**Significance:** These prediction models demonstrate the feasibility of early prognostication in *KCNQ2*-RD and support future prospective external validation. They enable more accurate individualised counselling by integrating clinical and genetic information readily available at time of genetic diagnosis and provide an objective foundation for early intervention planning and future precision medicine trial stratification.

## Introduction

*KCNQ2*-related disorders (*KCNQ2*-RD) represent a spectrum of early-onset neurological conditions caused by pathogenic variants in the *KCNQ2* gene (OMIM 602235), which encodes a voltage-gated potassium channel subunit (Kv7.2), essential for neuronal excitability (1). Kv7.2 co-assembles with Kv7.3 to generate the M-current, which counteracts cell depolarisation and dampens neuronal excitability when activated at subthreshold membrane potentials (2–6).

*KCNQ2*-RD are among the most common genetic causes of neonatal-onset epilepsy, representing 2-13% of genetic findings in individuals with epilepsy and neurodevelopmental disorders (7, 8). Associated phenotypes range from self-limited (familial) neonatal epilepsy (SeL(F)NE) to severe developmental and epileptic encephalopathy (DEE). *KCNQ2*-SeL(F)NE is characterised by self-limiting neonatal seizures, classically followed by normal neurodevelopment, whereas *KCNQ2*-DEE is associated with neonatal seizures and impairment of cognition, communication, and motor function (9–11).

Functional studies have linked the underlying molecular mechanisms of *KCNQ2* variants to clinical outcomes. Haploinsufficient variants, often truncating and dominantly inherited, cause moderate channel dysfunction through loss-of-function (LoF) mechanisms (up to 50% of M-current reduction) and are associated with the SeL(F)NE phenotype (2, 12). In contrast, severe DEE-associated variants arise *de novo*, cluster in functional hotspot regions, and cause more than 50% reduction in M-current density (13). These are most often missense variants exerting dominant-negative LoF effects (14). Lastly, rare missense variants associated with later-onset epilepsy and neurodevelopmental delay exhibit gain-of-function (GoF) effects (15, 16).

Yet, clinical experience and published data indicate a broader phenotypic spectrum (11, 17–19). Although (truncating) variants are expected to lead to a self-limiting course, developmental delay has been described in some individuals carrying SeL(F)NE-associated variants (20–22). Conversely, even carriers of recurrent DEE-associated missense variants can display substantial phenotypic heterogeneity, some achieving key milestones such as walking or verbal communication. Importantly, neonatal seizures occur in both groups, making early prediction of neurodevelopmental outcomes challenging. Moreover, functional characterisation is not available for all variants and is not always feasible in routine clinical practice. Notably, recent voltage-clamp studies conducted on recombinant human Kv7.2 channels showed only a modest correlation between the extent of channel dysfunction and clinical severity, suggesting that additional factors influence outcome (23). Thus, early prognostication based solely on genetic findings clearly has its limitations. Moreover, despite recent advances in genotype-phenotype atlases for neurodevelopmental disorders, such developmental trajectory mapping remains lacking for *KCNQ2*-RD, underscoring the need for granular stratification to better delineate developmental outcomes (24).

Machine learning algorithms can tackle complex prediction problems because they can integrate complex and incomplete datasets, and capture non-linear interactions between predictors (25, 26). In the context of *KCNQ2*-RD, previous computational efforts have focused primarily on variant interpretation. For example, the Bilbao Computational Biophysics (BCB) group developed an online tool to estimate the pathogenicity of missense variants. However, the model showed limited performance in distinguishing between phenotypic categories, namely SeL(F)NE and DEE, achieving a balanced accuracy of ∼67% (27). In addition, Xu et al. identified video-EEG features and variant location as predictors of poorer long-term outcome, based on multivariate ordinal logistic regression analysis (28). While valuable, these approaches do not provide granular, multi-domain developmental predictions.

Building on these prior efforts, we aimed to develop and internally validate machine-learning models that predict cognitive, language, and motor outcomes using clinical and genetic information available in early life. The models provide clinically applicable stratification of intellectual, verbal, and motor outcomes to (i) identify individuals at risk of developmental delay to optimise follow-up and multidisciplinary treatment and (ii) facilitate future precision therapy trials by stratifying individuals according to predicted outcome.

## Material and methods

### Study design and data collection

We conducted a multicentric, retrospective cohort study of individuals carrying a (likely) pathogenic variant in the *KCNQ2* gene. Participants were recruited through international epilepsy and genetic centres, *KCNQ2* patient organisations, the Network for Therapies in Rare Epilepsies (NETRE), the European Reference Network (ERN)-EpiCare Genetic Platform, and the IMPROVE registry. Enrolment took place between February 2021 and May 2026. Genetic and clinical data were collected via a standardised form completed by epileptologists, clinical geneticists, paediatric and adult neurologists.

The clinical data collected included epilepsy history, neurodevelopmental milestones, and outcomes. EEG classification was based on the terminology used in the original reports. Seizure types were classified according to detailed clinical descriptions when available, following the most recent ILAE guidelines (29). If detailed descriptions were not available, the classification provided by the treating physician was retained. Reported generalised tonic-clonic seizures in neonates were considered unlikely, as neonatal seizures typically have a focal onset; such cases were reclassified as sequential seizures in accordance with the neonatal ILAE classification (30).

### Inclusion and exclusion criteria

Only individuals with (likely) pathogenic variants in the *KCNQ2* gene, as classified by local diagnostic laboratories and confirmed by review according to ACMG criteria (31), were included. To ensure reliable neurodevelopmental outcomes, we only included participants aged three years or older at last follow-up. Moreover, individuals with known mosaic variants were excluded.

### *KCNQ2* variant definitions

According to the structural model of the Kv7.2 subunit described by Soldovieri et al. (6), variants were categorised based on their exon location and their presence within one of the four hotspots associated with the DEE phenotype, as defined by Millichap et al. (14)). These hotspots include: the S4 voltage-sensor, the pore loop, the proximal intracellular calmodulin-binding helix, and the distal calmodulin-binding helices.

Exons were grouped according to functional domains as follows: exons 1-2 correspond to the N-terminal domain; exons 3-5 to the voltage-sensing domain, including the S4 helix; exons 6-7 to the pore loop; exons 8-11 to the proximal C-terminal domain, including the proximal calmodulin-binding helix; and exons 12-17 to the distal C-terminal domain, including the distal calmodulin-binding helix.

Variants were classified as non-truncating (missense and in-frame deletions) or truncating (stopgain, frameshift, and splice-site variants).

### Neurodevelopmental outcome assessment and definition

Neurodevelopmental outcome was assessed by the treating physician using a predefined structured questionnaire, aligned with the DSM-5 intellectual disability (ID) severity domains (32). Outcomes were stratified into five ordered severity levels: normal intelligence (age-appropriate cognitive function), mild (minor developmental delays, largely independent), moderate (noticeable delays, requiring regular support), severe (major delays, dependent for most activities), and profound (extremely limited function, fully dependent). In a next step, while building models, outcome was grouped across normal intelligence, mild ID, and moderate to profound ID.

When available, standardised neuropsychological assessments were prioritised and used to assign severity categories according to predefined thresholds. Bayley cognitive scale or Vineland composite score ≥85 was considered normal, 70-84 mild, 55-69 moderate, 40-54 severe, and <40 profound. For IQ-based assessments (Wechsler scales), the corresponding ranges were: 50-70 mild, 35-49 moderate, 20-34 severe, and <20 profound. In the absence of formal testing, classification was derived from developmental history, milestone acquisition, and neurological examination.

All assessments were independently reviewed by two investigators (CM and EVB) based on clinical records and direct communication with treating clinicians and families, with discrepancies resolved by consensus. The cut-offs for ages of walking and of first words were based on the ages at which at least 95% of typically developing children achieve each milestone, according to the WHO Multicentre Growth Reference Study (33) and the Centers for Disease Control and Prevention guidelines (34). Accordingly, individuals were classified according to age at acquisition of first words (before, after 16 months, or never achieved), and of walking (before, after 18 months, or never achieved).

We recognise that a small minority of children may acquire first words or walking after age of inclusion, set at a minimum of three years. To balance methodological consistency with clinical realism, we applied the following rule: if a child had not achieved a milestone by age three and no later documentation was available, the milestone was classified as “never achieved” at last follow-up; if subsequent records confirmed later acquisition, the classification was updated accordingly. Across the full cohort of 277 individuals, only seven children (7/277, 3%) produced first words after 3 years of age, and eight (8/277, 3%) achieved independent walking after 3 years of age. Given the very low frequency of such late attainment and the need to maximise sample inclusion for model development, the three-year threshold was considered reasonable.

## Statistical analysis

Descriptive statistics were used to summarise demographics and the distribution of clinical and genetic variables, grouped by outcome category. Categorical variables were presented as frequencies and percentages; continuous variables were presented as medians with interquartile ranges (IQR).

For each developmental outcome (ID, age of first words and age of walking), random forest models were built. Candidate neonatal predictors were selected a priori based on biological plausibility, published literature and expert clinical consensus (SW and MM), and were extracted from the collected clinical and genetic patient files. Among 15 clinical and genetic parameters, the ones with more than 50% data availability were retained. The same 10 predictors were used across all models to ensure clinical consistency and applicability (Tables 1, S2 and S3, Fig. S1). Parameters not retained for model building due to insufficient data were birth length, weight, and head circumference, the presence of congenital malformations, and brain MRI at seizure onset. Their distribution is represented in Table S1. As exploratory analyses, univariable tests were performed to assess associations between these non-retained parameters and neurodevelopmental outcome. Chi-square tests were performed for categorical variables, ANOVA tests for continuous variables with a normal distribution, and Kruskal-Wallis tests for continuous variables with a non-normal distribution.

Next, the dataset was randomly partitioned into training (70%) and validation (30%) cohorts using stratified sampling to preserve outcome class proportions across datasets. To prevent data leakage, we ensured that no variant occurred in both the training and validation set.

Three random forest models were developed using an identical set of ten clinical and genetic input features. Models were implemented using the Breiman random forest algorithm as available in the R package randomForest, while the caret package was used for data partitioning, stratified cross-validation, and performance assessment (35). The number of variables randomly sampled at each split (mtry) was optimised using the tuneRF procedure based on minimisation of out-of-bag classification error. Missing values within the training set were handled using the na.roughfix function from the randomForest package, replacing categorical variables with the most frequent category. Missing data are summarised in table S4. To address class imbalance, inverse-frequency class weights (classwt) were incorporated during model training.

Predictive performance was evaluated using stratified 5-fold cross-validation. Mean accuracy and Cohen’s kappa values were calculated across folds, with empirical 95% confidence intervals derived from fold-wise variability. Final model performance was subsequently assessed on the independent hold-out test cohort using accuracy, macro-averaged F1-score, Cohen’s kappa, and one-vs-rest receiver operating characteristic (ROC) analyses with area under the curve (AUC) estimates for each outcome class. Variable importance was assessed using mean decrease in Gini impurity and mean decrease in accuracy. Input feature effects and model interpretability were further explored post hoc using partial dependence plots (PDPs) and SHapley Additive exPlanations (SHAP) values.

As exploratory analyses, post hoc chi-square tests were performed to assess associations between early clinical/genetic variables and neurodevelopmental outcome severity. Chi-square tests of independence were applied to categorical variables across outcome groups, followed by examination of standardised residuals to identify category-outcome combinations contributing most strongly to significant associations (Table 1).

**Table 1.** Demographics and summary according to degree of neurodevelopmental delay. Missing data are not displayed. ^1^Results of chi-square tests for categorical variables. The reported *P* values correspond to global chi-square tests of independence across all groups. To identify which category-severity combinations contributed to this association, cells with |standardised residual| ≥ 2 are highlighted (□: over-represented, ■: under-represented). IED: interictal epileptiform discharge, TV: truncating variant.

| Variable | Groups |  |  | P value<sup>1</sup> |
| --- | --- | --- | --- | --- |
| Neurodevelopmental delay | No | Mild | Moderate to profound |  |
| **De novo** |  |  |  | <0.001 |
| De novo (N=133) | 17 (20%)■ | 19 (58%) | 96 (95%)□ |  |
| Inherited (N=89) | 70 (81%)□ | 14 (42%) | 5 (5%)■ |  |
| **Exon** |  |  |  | <0.001 |
| 1-2 (N=14) | 10 (9%)□ | 2 (6%) | 2 (2%) |  |
| 3-5 (N=79) | 15 (13%)■ | 15 (42%) | 49 (41%) |  |
| 6-7 (N=54) | 6 (5%)■ | 4 (11%) | 44 (37%)□ |  |
| 8-11 (N=18) | 12 (10%)□ | 1 (3%) | 5 (4%) |  |
| 12-17 (N=43) | 20 (17%) | 4 (11%) | 18 (15%) |  |
| TV (N=69) | 55 (47%)□ | 10 (28%) | 4 (1%)■ |  |
| **Hotspot** |  |  |  | <0.001 |
| Non hotspot (N=55) | 35 (30%)□ | 7 (19%) | 13 (10%)■ |  |
| Hotspot (N=153) | 28 (24%)■ | 19 (53%) | 105 (88%)□ |  |
| TV (N=69) | 55 (47%)□ | 10 (28%) | 4 (2%)■ |  |
| **Seizure onset age** |  |  |  | <0.001 |
| First day (N=70) | 10 (9%)■ | 9 (25%) | 51 (44%) |  |
| 2<sup>nd</sup> day – 1 month (N=160) | 80 (69%) | 24 (67%) | 56 (48%) |  |
| After 1 month / never (N=40) | 26 (22%) | 3 (8%) | 10 (9%) |  |
| **Seizure onset type** |  |  |  | <0.001 |
| Sequential (N=123) | 71 (64%)□ | 18 (53%) | 34 (30%)■ |  |
| Tonic (N=91) | 15 (14%)■ | 13 (38%) | 63 (56%)□ |  |
| Other / no seizures (N=45) | 25 (23%) | 3 (9%) | 16 (14%) |  |
| **EEG at seizure onset** |  |  |  | <0.001 |
| Normal (N=39) | 28 (53%)□ | 3 (12%)■ | 8 (8%)■ |  |
| Burst suppression (N=42) | 3 (6%)■ | 3 (12%) | 36 (37%)□ |  |
| (Multi)focal IEDs with/without background slowing (N=95) | 22 (42%)■ | 20 (77%) | 53 (55%) |  |
| **Frequency of seizures at onset** |  |  |  | 0.001 |
| Multiple per day (N=167) | 62 (61%)■ | 21 (70%) | 84 (84%)□ |  |
| Less than multiple per day (N=65) | 39 (39%)□ | 9 (30%) | 16 (16%)■ |  |
| **Hypotonia at birth** |  |  |  | <0.001 |
| No (N=156) | 110 (94%)□ | 22 (67%) | 24 (22%)■ |  |
| Yes (N=106) | 7 (6%)■ | 11 (33%) | 87 (78%)□ |  |
| **Prematurity** |  |  |  | 0.682 |
| Present (N=26) | 7 (7%) | 3 (10%) | 15 (16%) |  |
| Absent (N=200) | 96 (93%) | 26 (90%) | 78 (84%) |  |
| **Sex** |  |  |  | 0.701 |
| Female (N=125) | 58 (53%) | 10 (30%) | 57 (56%) |  |
| Male (N=120) | 51 (47%) | 23 (70%) | 45 (44%) |  |

As previously demonstrated in this cohort, individuals with *KCNQ2*-DEE showed more favourable outcomes when sodium channel blockers (SCBs) were initiated early (≤1 month of age) (36). The SCBs used in this cohort were carbamazepine, oxcarbazepine, lacosamide, lamotrigine, and phenytoin. To mitigate the confounding effect of age at SCB initiation on neurodevelopmental outcome, we adjusted outcome measures using a linear regression-based standardisation approach on the training dataset. Neurodevelopmental outcome was regressed on age at SCB initiation, and for each individual, the observed outcome was adjusted to the value expected at the cohort’s mean age of treatment initiation. This SCB-adjusted continuous severity score was subsequently used as the dependent variable in the random forest regression model. Model performance was evaluated using the root mean squared error (RMSE), mean absolute error (MAE), and coefficient of determination (R²). As this sensitivity analysis did not materially change individual severity categorisation and showed only moderate explanatory performance (R² = 0.485), primary analyses were conducted using the original categorical outcome model, which provides more clinically interpretable stratification of neurodevelopmental severity.

A summary workflow of the methodology is represented in Fig. 1. We adhered to established guidelines for the development and reporting of prediction models using artificial intelligence (37). All analyses were performed using R (version 4.5.1).

**Figure 1.**
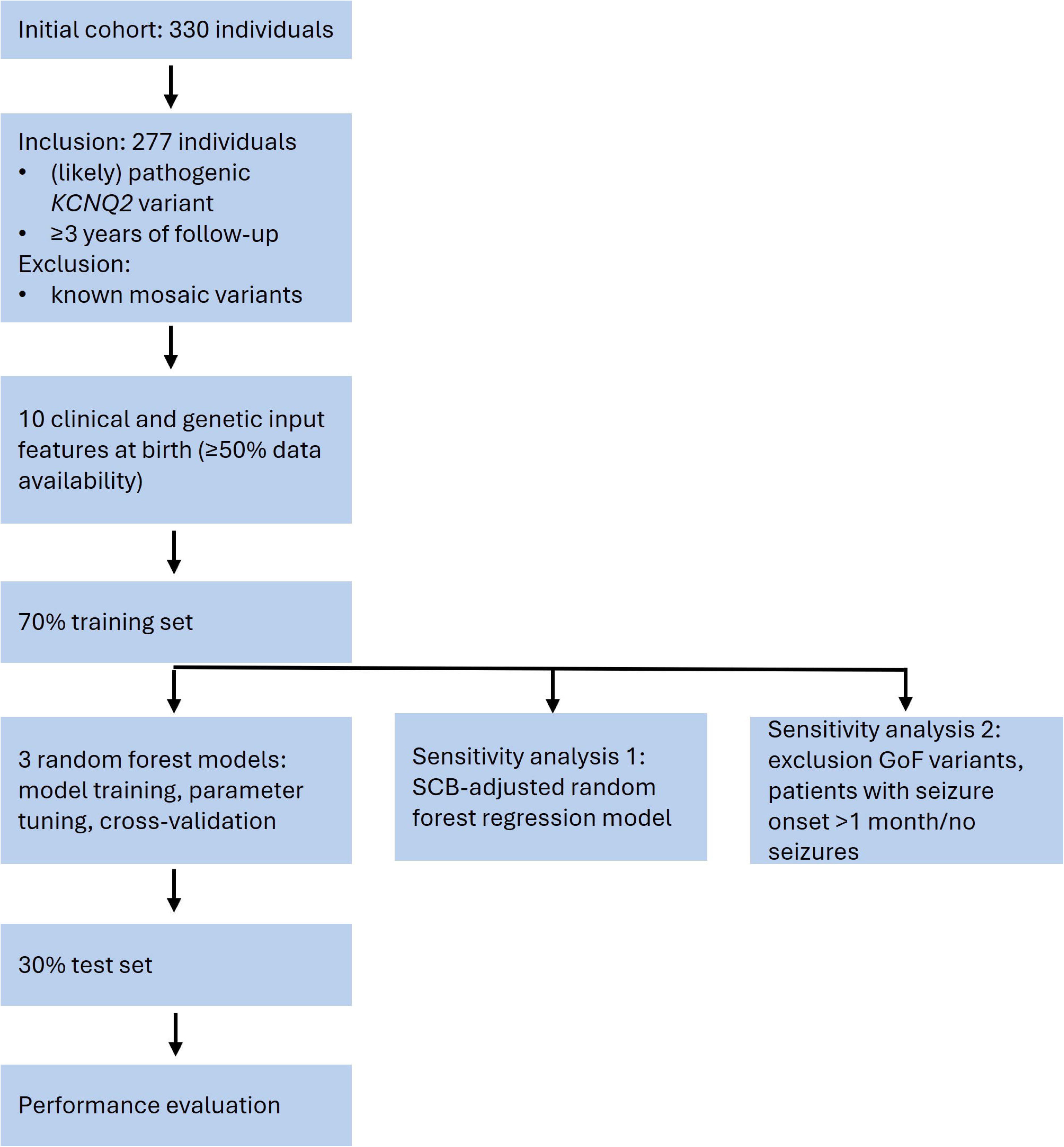
Workflow of the construction of random forest models to predict neurodevelopmental delay, age of talking, and of walking in *KCNQ2*-RD. GoF: gain-of-function, SCB: sodium channel blocker.

### Standard Protocol Approvals, Registrations, and Patient Consents

The study was approved by the Human Research Ethics Committee of the University Hospital of Antwerp (Belgium, reference 20/50/683) and the Comité de Protection des Personnes Sud-Est III (France, IDRCB 2020-A01363-36). Reporting follows the STROBE (Strengthening the Reporting of Observational Studies in Epidemiology) guidelines. Informed consent for participation and publication was obtained from all participants, their parents or legal guardians in accordance with the Declaration of Helsinki.

## Data Acces and Availability

The first authors (EVB and CM) and the last author (SW) have full access to all study data, take responsibility for the integrity of the data and the accuracy of the analyses, and have the right to publish the data independently of any sponsor. Anonymised data not published within this article will be made available by request from any qualified investigator.

## Results

### Participant characteristics

Clinical and genetic data were collected from 330 individuals across 23 countries. Following evaluation of the genetic variants, four variants of unknown significance and six mosaic variants were excluded. A further 43 individuals were excluded due to an age at last follow-up lower than three years.

Demographics and distribution of clinical and genetic variables at onset are grouped by outcome category (Tables 1, S1, S2 and S3). A visual representation of the distribution of variables according to outcome is represented in Fig. S1.

The study population comprised 277 individuals, including 125 females, 120 males, while sex information was unavailable for 32 individuals. The median age at last follow-up was 105 months (IQR 55-218). The median age at seizure onset was two days (IQR 2-4). Ninety-two percent (254/277) of individuals achieved seizure freedom at last follow-up, with a median age of seizure offset of 4 months (IQR 1-12), in line with recent cohort and natural history studies (2, 38).

The median age at which first words were achieved was 22 months (IQR 12-100), and the median age at independent walking was 19 months (IQR 13-100). In terms of neurodevelopmental outcomes, 119 individuals (43%) exhibited normal development, 38 (14%) had mild ID, and 120 (43%) showed moderate to profound ID. Across the 158 individuals with some degree of ID, formal neuropsychological assessment was available for 68 (43%). The remainder were classified using predefined clinical criteria. Fifty percent (138/277) of individuals had been treated with SCBs and 24% (66/277) initiated the treatment before one month of age. Median age of start of SCB was 32 days (IQR 7-267).

### Random forest analysis

Seven early clinical parameters were used to train the models: the presence of hypotonia at birth, EEG characteristics performed within three days after seizure onset (normal, burst suppression or other abnormal EEG), the age at seizure onset (first day of life, second day to one month of life or after one month/never), the type of seizures at onset (sequential seizures, tonic seizures or other/no seizures) and their frequency (multiple per day or less frequent), the presence of prematurity and the sex; along with three genetic predictors: the exon in which the variant is located, taking into account the type of variant (truncating or non-truncating), the location within one of the known hotspots for *KCNQ2*-DEE, and the presence of a *de novo* variant (tables 1, S2 and S3, Fig. S1). Other abnormal EEGs included focal or multifocal interictal epileptiform discharges (IEDs), non-physiological background slowing, and hypsarrhythmia.

First, three dichotomous classification models were developed to predict (i) neurodevelopmental outcome (normal intelligence versus mild-to-profound ID) (ii) language acquisition (achieved versus never achieved), and (iii) attainment of independent walking (achieved versus never achieved). The accuracies on the test set were 0.83, 0.83, and 0.86, and the corresponding macro F1 scores were 0.89, 0.85, and 0.92, respectively. To enable more granular prognostication for clinical counselling and more precise outcome-based stratification for future clinical trials, three-category classification models were subsequently built to predict (i) neurodevelopmental outcome (normal intelligence vs. mild ID vs. moderate-to-profound ID), (ii) age at first words (<16 months vs. >16 months vs. never achieved), and (iii) age at independent walking (<18 months vs. >18 months vs. never achieved). These models achieved accuracies of 0.79, 0.70, 0.71, and macro F1 scores of 0.85, 0.82, and 0.83, respectively. The performance of the models is summarised in table 2. The one-vs-rest Receiver Operating Characteristic (ROC) curves and confusion matrices of the models are represented in Fig. 3.

**Table 2.** Predictive performance of random forest models for neurodevelopmental outcomes in *KCNQ2*-RD. Model performance was evaluated using stratified 5-fold cross-validation and independent hold-out test set validation. Cross-validation results are presented as mean estimates with empirical 95% confidence intervals. ID: intellectual disability.

| Model | Cross-validation |  | Test set |  |  |
| --- | --- | --- | --- | --- | --- |
|  | Cohen's Kappa | Accuracy | Cohen's Kappa | Macro F1 score | Accuracy |
| **Two-category classification** |  |  |  |  |  |
| Neurodevelopmental delay (normal intelligence vs. mild-profound ID) | 0.74 (0.65-0.82) | 0.89 (0.85-0.94) | 0.76 | 0.89 | 0.83 |
| Acquisition of words (achieved vs. never achieved) | 0.65 (0.59-0.74) | 0.85 (0.80-0.92) | 0.64 | 0.85 | 0.83 |
| Acquisition of walking (achieved vs. never achieved) | 0.66 (0.60-0.78) | 0.84 (0.79-0.87) | 0.64 | 0.92 | 0.86 |
| **Three-category classification** |  |  |  |  |  |
| Neurodevelopmental delay (normal intelligence vs. mild vs. moderate-profound ID) | 0.52 (0.49-0.75) | 0.79 (0.72-0.86) | 0.60 | 0.85 | 0.79 |
| Age of first words ($\leq 16$ months vs. $> 16$ months vs. never achieved) | 0.51 (0.40-0.71) | 0.69 (0.65-0.72) | 0.49 | 0.82 | 0.70 |
| Age of walking ($\leq 18$ months vs. $> 18$ months vs. never achieved) | 0.55 (0.45-0.63) | 0.75 (0.65-0.83) | 0.51 | 0.83 | 0.71 |

Variable importance analysis identified seven variables that consistently ranked highest across the models for neurodevelopmental outcome: presence of a *de novo* variant, exon group, location within a *KCNQ2* DEE-hotspot, hypotonia at birth, type of seizure at onset, EEG pattern, and age at seizure onset (Fig. 2.).

**Figure 2.**
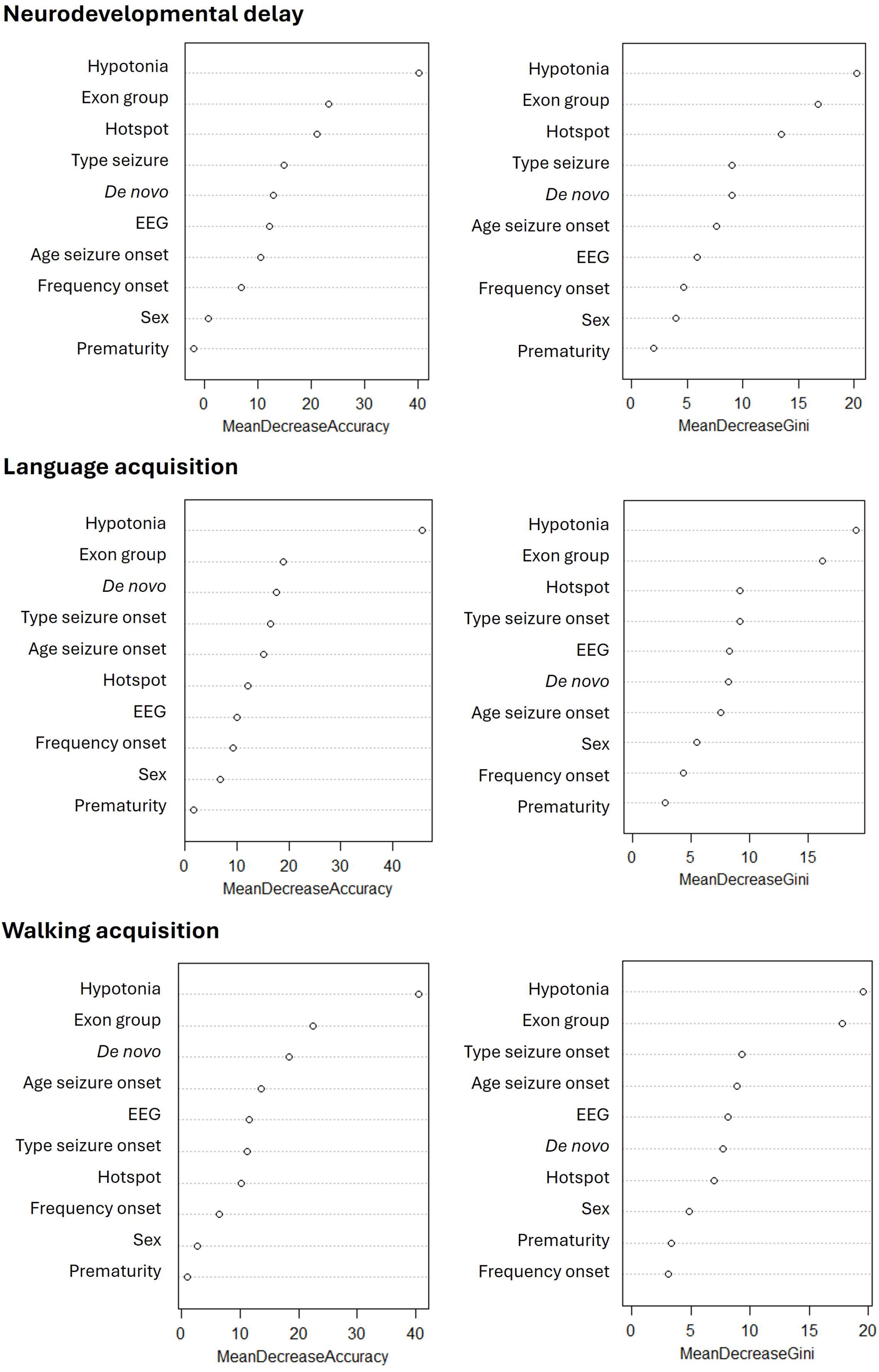
Variable importance in Random Forest models. Importance of each predictor is expressed as the mean decrease in accuracy (MDA) and mean decrease in Gini index (MDG) across all trees in the model.

**Figure 3.**
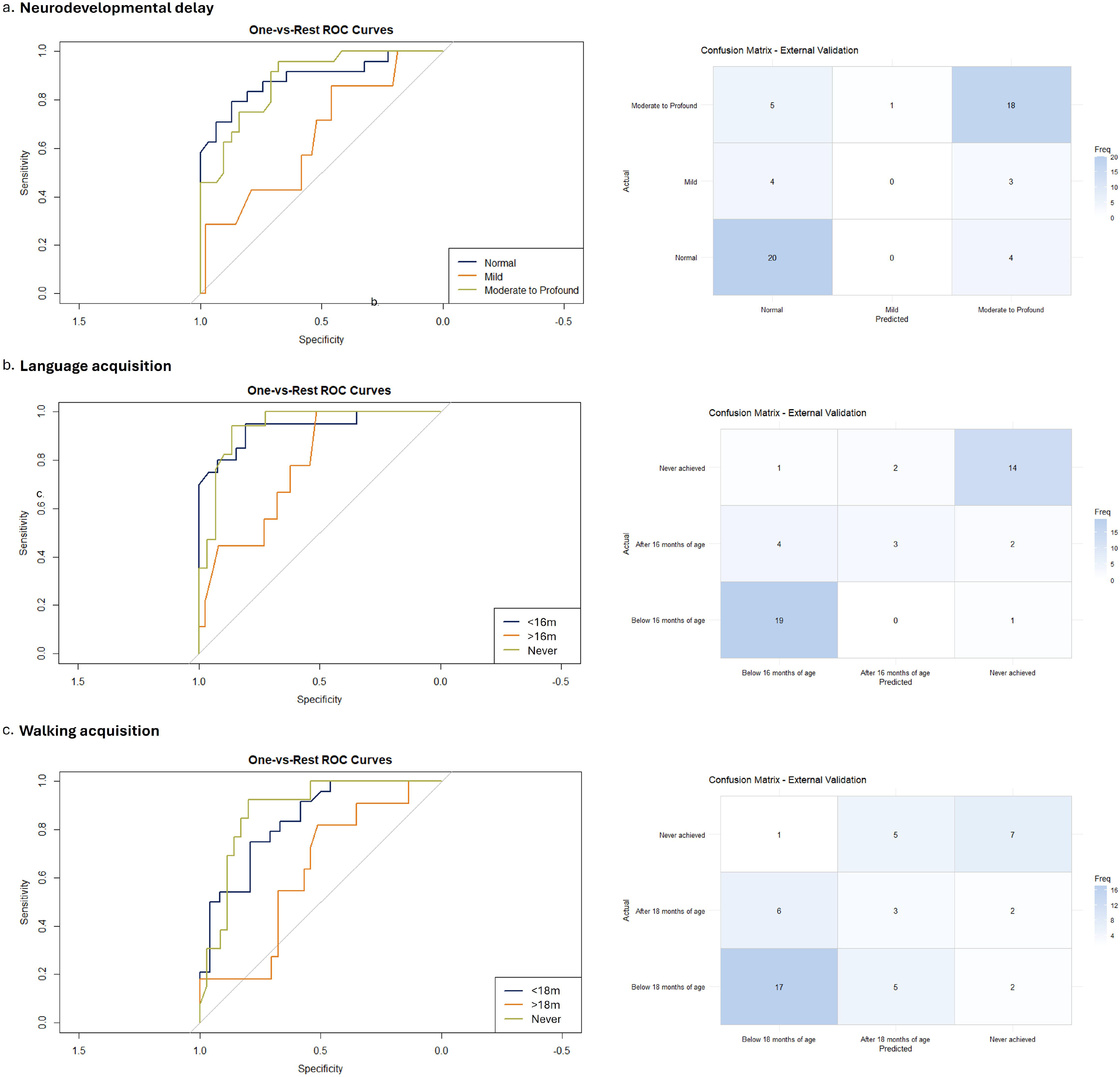
One-vs-rest Receiver Operating Characteristic (ROC) curves and confusion matrix for the random forest model predicting neurodevelopmental outcomes. Area under the ROC curve (AUC) was 0.89 for normal, 0.65 for mild, and 0.88 for moderate to profound delay (**3a**). One-vs-rest ROC curves and confusion matrix for predicting age of first words. AUC was 0.94 for acquisition before 16 months, 0.77 for acquisition after 16 months, and 0.94 for words never achieved (**3b**). One-vs-rest Receiver ROC curves for predicting age of walking. AUC was 0.89 for walking before 18 months, 0.75 for walking after 18 months, and 0.88 for walking never achieved (**3c**).

For neurodevelopmental outcome, partial dependency analysis revealed that hypotonia at birth (predicted value 0.70), a *de novo* variant (0.81), seizure onset within the first day of life (0.62), variants in exon group 6-7 (0.83), localisation within a DEE hotspot (0.59), multiple seizures per day at onset (0.55), tonic seizures at onset (0.59) and a burst-suppression EEG pattern (0.54) were associated with worse cognitive outcome. PDPs for each outcome can be found in the Supporting Information (Fig. S2-S4).

### Case studies

Within our cohort, six individuals carried the recurrent p.(Arg213Trp) variant, known to cause dominant-negative LoF effects, associated with *KCNQ2*-DEE (13). Four of these six individuals presented with moderate to profound ID, all of whom were correctly classified by the random forest model (Fig. S5). The remaining two individuals exhibited a milder phenotype with normal intelligence. Notably, among the six individuals carrying this variant, only these two individuals received early treatment with SCBs. One of these individuals (Fig. 4a) was correctly predicted as having a normal outcome by the model. In contrast, the second individual (Fig. 4b) was misclassified as having moderate to profound ID. SHAP analysis indicated that the feature contributing most strongly to the prediction of normal intelligence and moderate to profound ID in these two respective individuals was the absence or presence of neonatal hypotonia (phi: 0.09 for the first and 0.18 for the second individual), consistent with its high importance in the mean decrease Gini and mean decrease accuracy variable-importance plots.

**Figure 4.**
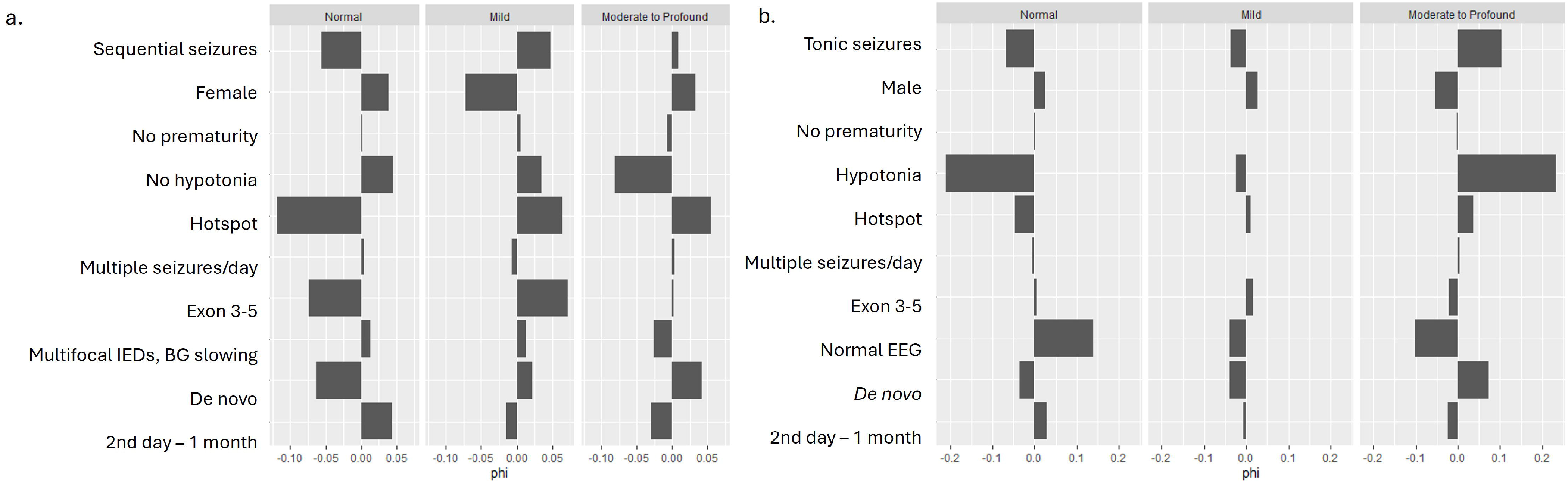
Case study of individuals carrying the recurrent *KCNQ2*-DEE associated variant p.Arg213Trp. Four patients had, as expected, moderate to profound intellectual disability (ID), and were correctly predicted by the model, while two had normal cognitive outcome. The SHapley Additive exPlanations (SHAP) analysis of the two individuals with normal cognitive outcome is represented. These two were also the only to receive early sodium channel blockers (SCBs). The first was correctly predicted by the model, mainly based on the absence of hypotonia at birth (**4a**). The second individual was predicted as moderate to severe intellectual disability, mainly based on the presence of hypotonia at birth, but with possible influence of SCB (**4b**).

Among the individuals in our cohort misclassified for neurodevelopmental outcome, seven had better-than-predicted outcomes, and five of them received SCBs in the first month of life. Next, a total of seven individuals with neonatal hypotonia had normal cognitive outcome, six of them received SCBs in the first month of life. Among individuals with worse-than-predicted outcome, one developed Infantile Epileptic Spasm Syndrome (after five months of age), characterised by epileptic spasms, hypsarrhythmia on EEG, and clinical regression. He had a GoF p.(Arg144Gln) variant, not located within a known DEE-hotspot, and did not present with hypotonia at birth, nor neonatal seizures.

Finally, as individuals with known mosaic variants were excluded from the primary cohort, a post-hoc exploratory SHAP analysis was performed in one individual carrying the recurrent p.(Arg213Trp) variant with confirmed mosaicism, who presented with a milder-than-expected phenotype and normal intelligence. The model correctly predicted a normal neurodevelopmental outcome in this individual.

## Discussion

We developed machine-learning models capable of predicting neurodevelopmental outcomes in individuals with *KCNQ2*-RD using routinely available neonatal clinical and genetic data. Our overarching aim was to support early prognostic counselling, enable timely identification of infants at risk of adverse developmental trajectories, and facilitate individual stratification for future precision-medicine trials.

In keeping with beside clinical decision-making, we first implemented binary classifiers and then extended to three-level models. The binary approach (normal vs. mild-profound ID; acquisition or not of first words and independent walking) yielded the strongest validation performance and therefore serves as a robust first step for early risk stratification. Although the three-class models showed slightly less performance (macro F1 scores diminished by 3-9% and accuracies by 4-15%), they offer clinically meaningful discrimination between levels of impairment (normal, mild, moderate-profound ID; first words ≤16 months, >16 months, never; walking ≤18 months, >18 months, never). This added granularity is relevant for counselling families about likely support needs, and it is particularly useful for trial stratification.

A pragmatic future application would be a two-step approach in which binary classification first identifies individuals at increased risk to guide counselling, developmental surveillance, and referral pathways, followed by multiclass prediction to provide more detailed prognostic information and support stratification for early-intervention studies and future precision-medicine trials. This approach balances predictive performance with the granularity required for real-world decisions.

Across model training, validation, and post-hoc interpretability analyses, including variable importance measures, PDPs, SHAP analyses, and post-hoc univariable testing; hypotonia at birth, seizure onset within the first day of life, frequent and tonic neonatal seizures, burst-suppression EEG pattern at onset, de novo variants, and variants located within exons 6-7 consistently emerged as key predictors of adverse neurodevelopmental outcome in *KCNQ2*-RD.

Neonatal hypotonia consistently emerged among the most influential predictors across all analyses. This is consistent with clinical practice and previous literature linking hypotonia at birth with poorer neurodevelopmental trajectories in neonatal epilepsies, emphasizing the importance of early and detailed clinical assessment (39).

As reported previously, localisation of variants also contributed to outcome stratification. Variants in the pore-forming regions (exons 6-7) were associated with the most severe phenotypes, while changes in the N-terminal domain (exons 1-2), and in the proximal C-terminal domain (exons 8-11) showed more favourable outcomes (28, 40). At the same time, outcome variability persists even among recurrent DEE-associated missense variants, indicating that genetic data alone are insufficient for reliable early prognostication. Together with the modest genotype-phenotype coupling reported in functional studies (23), these observations support a multifactorial prediction framework that integrates clinical and genetic features captured in the neonatal period.

Regarding EEG features, burst suppression was associated with the highest predicted probability of moderate to profound ID, followed by another abnormal EEG, whereas a normal EEG was more associated with normal intelligence (Fig. S2).

Truncating variants are well-known to be associated with a milder phenotype (2), in line with our findings. As expected, in our cohort, only 2% of individuals with truncating variants had moderate to profound ID.

Inherent to a retrospective study in rare disorders, this study has some limitations. First, the relative limited sample size and class imbalance likely contribute to lower performance in the intermediate categories, as reflected by one-vs-rest AUC values and the confusion matrices. Next, the retrospective design may further have led to unmeasured confounding or incomplete capture of predictors and outcomes. Neurodevelopmental outcomes were assessed using multiple instruments, and in some cases approximated from developmental history, milestones, and physical examination when formal testing was unavailable. This potential variability and misclassification, particularly in borderline cases and across middle categories, could affect model training and predictive accuracy, although independent review by CM and EVB was performed to mitigate this.

A further limitation relates to EEG classification, which was based on routine clinical reports rather than centralised review of the original recordings. This approach may have introduced inter-rater variability and precluded the application of standardised neurophysiological definitions, particularly for burst-suppression for which definitions vary in the literature. Moreover, because truly normal EEGs (without any epileptiform abnormalities) are not so common in *KCNQ2*-RD, the “normal” category reflected the reporting clinician’s overall interpretation of the background and may have included a certain degree of abnormalities. It is also possible that the intermediate category of other abnormal EEGs may include both severe tracings (with few physiological patterns, an alteration in sleep organisation) and tracings with milder abnormalities, in which the background is well preserved but spikes are more abundant (typically predominant in the bilateral central regions). This could explain a less discriminatory role of EEG in distinguishing between favourable and unfavourable outcomes. Finally, only the EEG pattern at seizure onset was considered. As EEG abnormalities in *KCNQ2*-RD evolve dynamically during the neonatal period, often in parallel with treatment initiation, longitudinal assessment of EEG evolution may provide greater prognostic value. Future studies incorporating centralised EEG review and serial neurophysiological assessment, particularly following SCB initiation, may further improve prognostic accuracy.

To maximise the cohort size and optimise the applicability of our prediction model, we also included 17 asymptomatic carriers of a pathogenic *KCNQ2* variant (17/277, 6%) who were diagnosed because family members had *KCNQ2*-related epilepsy, and 12 individuals (12/277, 4%) with GoF variants and later onset or absent epilepsy. Although such individuals would not be identified clinically in the neonatal period, their inclusion increased both the sample size and the representation of the full phenotypic spectrum of *KCNQ2*-RD. In our model, the category seizure onset after one month, capturing most GoF variants, was associated with moderate-profound ID (predicted probability of 0.54). However, one individual with worse-than-predicted outcome carried a GoF variant, highlighting the need to increase the number of individuals for future studies on this variant type. A sensitivity analysis was performed in a restricted subgroup excluding individuals with late seizure onset (>1 month), absence of seizures, and GoF variants, to assess the robustness of model performance in a more closely reflecting the target clinical population. In this restricted cohort, the model demonstrated good predictive performance on the test set, with an accuracy of 0.80, a Cohen’s kappa of 0.64, and a macro-averaged F1-score of 0.89 (Fig. S6). This supports the potential translational relevance of the model.

By design, our model estimates the expected neurodevelopmental trajectory from baseline features available at birth, and does not account for post-baseline interventions that may modify outcomes, such as early initiation of SCBs, intensity and timing of rehabilitation, speech-language therapy. It is therefore intended for early prognostic counselling and risk stratification. Although early SCB treatment has previously been associated with improved outcomes (36), a sensitivity analysis adjusting for SCB initiation did not materially alter individual severity classification. This suggests that baseline clinical and genetic features remain the main drivers of prognostic stratification in this dataset, while not excluding meaningful treatment effects at the individual level. Larger prospective cohorts incorporating time-varying treatment exposures will be required to develop dynamically updated prognostic models.

We are currently developing an online prediction tool within the KCNQ2 Portal (https://kcnq2-portal.lalresearchgroup.org/) to facilitate the clinical implementation of these models. A direct link to the prediction tool will be provided in the revised version of the manuscript.

## Conclusion

In our cohort of individuals with *KCNQ2*-RD, we developed and internally validated machine learning models to predict neurodevelopmental outcome based on neonatal features. Individual-level examples illustrate how incorporating a broader range of predictors allows for more finely tuned prediction of neurodevelopmental trajectories. Overall, this prediction tool may aid in selecting individuals for targeted therapies and more intensive early management, and provides a foundation for future, intervention-aware validation and deployment in clinical practice.

## Appendix

*KCNQ2* study group: Lucia Abela, Laura Abraira del Fresno, Nicholas Allen, Jürgen Althaus, Amaia Lasa Aranzasti, Stéphane Auvin, Susann Badmann, Claire Bar, Nina Barisic, Tania Barragán Arévalo, Andrea Berger, Luca Bergonzini, Tobias Bartolomaeus, Ernie Bongers, Beatrice Desnous, Rebecca Gembicki, Britta Hanker, Charlotte Thiels, Christine Makowski, Elisabeth Cats, Stephanie Colling, Matthias De Wachter, Anne De Saint-Martin, Geoffroy Delplancq, Blandine Dozieres, Nathalie Dorison, Francesca Madia, Dagmar Wieczorek, Caroline Hachon-Le Camus, Johanneke Harteman, Katrine Marie Harries Johannesen, Benedicte Heron, Kathleen Gorman, Janina Gburek-Augustat, Michael Gößler, Joerg Klepper, Ilona Krey, Mathieu Kuchenbauch, Lieven Lagae, Amaia Lasa Aranzasti, Anne Lepine, Jeremie Lefranc, Ana Isabel Lopes Vilan, Oliver Maier, Isabella Mauro, Niamh McSweeney, Cyril Mignot, Rikke Steensbjerre Møller, Silvia Adriana Napuri Peirano, Mary O’Regan, Paloma Parra, Caroline Perriard, Elena Pavlidis, Pegoraro Veronica, Filip Roelens, Angelo Russo, Antonio Gil-Nagel, Katrien Jansen, Silvia Jorge, Eulalia Turon Viñas, Giulia Ventura, Nathalie Villeneuve, Laurent Villard, Dorothée Ville, Marketa Vlckova, Patrick Van Bogaert, Katrien Vanrykel, Nienke Verbeek, Helene Verhelst, Pasquale Striano, Carlotta Stipa, Mariël Teunissen, Carmen Laura Trujillano Lidon, Pia Zacher, Jana Zarubova

