## Supplemental Material for "Early clinical prediction of neurodevelopmental outcome in *KCNQ2*-related disorders"

### Supporting Information

| Variable | Groups | | | *P* value[1] |
|---|---|---|---|---|
| **Neurodevelopmental delay** | **No** | **Mild** | **Moderate to profound** | |
| Birth length (cm) | 50 (48.5-54) | 52 (50-54) | 50 (47.5-51) | 0.507 |
| Birth weight (kg) | 3.450 (3.170-3.750) | 3.623 (3.416-3.978) | 3.220 (2.820-3.500) | 0.643 |
| Birth head circumference (cm) | 34 (33.5-35.5) | 35 (35-35) | 35 (33.5-36.5) | 0.441 |
| **Congenital malformations** | | | | 0.868 |
| Present (*N*=10) | 3 (3%) | 0 (0%) | 7 (10%) | |
| Absent (*N*=184) | 95 (97%) | 24 (100%) | 67 (90%) | |
| **Prematurity** | | | | 0.682 |
| Present (*N*=25) | 6 (6%) | 3 (11%) | 15 (16%) | |
| Absent (*N*=196) | 95 (94%) | 24 (89%) | 77 (84%) | |
| **Sex** | | | | 0.701 |
| Male (*N*=115) | 48 (45%) | 22 (71%) | 44 (44%) | |
| Female (N=124) | 59 (55%) | 9 (29%) | 56 (56%) | |
| **Brain MRI at seizure onset** | | | | 0.333 |
| Normal (*N*=92) | 25 (83%) | 12 (86%) | 54 (76%) | |
| Abnormal (*N*=24) | 5 (17%) | 2 (14%) | 17 (24%) | |

**Table S1.** Potential clinical predictors not retained for the random forest models. Continuous variables are represented as median with interquartile range. Missing data are not displayed. [1]Results of chi-square tests for categorical variables and ANOVA tests for continuous variables with a normal distribution and Kruskal-Wallis tests for continuous variables with a non-normal distribution.

| Variable | Groups | | | *P* value[1] |
|---|---|---|---|---|
| **Age of first words** | **Below 16 months** | **After 16 months** | **Never achieved** | |
| ***De novo*** | | | | <0.001 |
| *De novo* (*N*=133) | 19 (26%) | 29 (73%) | 69 (99%) | |
| Inherited (*N*=89) | 55 (74%) | 11 (28%) | 1 (1%) | |
| **Exon** | | | | <0.001 |
| 1-2 (*N*=14) | 7 (7%) | 0 (0%) | 2 (2%) | |
| 3-5 (*N*=79) | 13 (13%) | 18 (40%) | 32 (39%) | |
| 6-7 (*N*=54) | 8 (8%) | 4 (9%) | 38 (45%) | |
| 8-11 (*N*=18) | 10 (10%) | 4 (9%) | 1 (1%) | |
| 12-17 (*N*=43) | 15 (16%) | 12 (27%) | 8 (9%) | |
| TV (*N*=69) | 44 (45%) | 7 (16%) | 3 (3%) | |
| **Hotspot** | | | | <0.001 |
| Non hotspot (*N*=55) | 26 (27%) | 6 (13%) | 7 (8%) | |
| Hotspot (*N*=153) | 27 (28%) | 32 (71%) | 74 (89%) | |
| TV (*N*=69) | 44 (45%) | 7 (16%) | 3 (3%) | |
| **Seizure onset age** | | | | 0.020 |
| First day (*N*=70) | 11 (12%) | 19 (43%) | 32 (41%) | |
| 2nd day – 1 month (*N*=160) | 63 (66%) | 20 (46%) | 41 (52%) | |
| After 1 month / never (*N*=40) | 22 (23%) | 5 (11%) | 6 (8%) | |
| **Seizure onset type** | | | | 0.001 |
| Sequential (*N*=123) | 62 (65%) | 18 (42%) | 21 (28%) | |
| Tonic (*N*=91) | 17 (18%) | 21 (49%) | 42 (55%) | |
| Other / no seizures (*N*=45) | 17 (18%) | 4 (9%) | 13 (17%) | |
| **EEG at seizure onset** | | | | <0.001 |
| Normal (*N*=39) | 21 (47%) | 9 (24%) | 4 (6%) | |
| Burst suppression (*N*=42) | 5 (11%) | 7 (18%) | 26 (40%) | |
| (Multi)focal IEDs with/without background slowing (*N*=95) | 19 (42%) | 22 (58%) | 35 (54%) | |
| **Frequency of seizures at onset** | | | | 0.001 |
| Multiple per day (*N*=167) | 58 (65%) | 24 (67%) | 55 (86%) | |
| Less than multiple per day (*N*=65) | 31 (35%) | 12 (33%) | 9 (14%) | |
| **Hypotonia at birth** | | | | <0.001 |
| No (*N*=156) | 86 (91%) | 20 (48%) | 11 (14%) | |
| Yes (*N*=106) | 9 (10%) | 22 (52%) | 66 (86%) | |
| **Prematurity** | | | | 0.892 |
| Present (*N*=26) | 7 (8%) | 6 (18%) | 9 (15%) | |
| Absent (*N*=200) | 81 (92%) | 28 (82%) | 53 (86%) | |
| **Sex** | | | | 0.754 |
| Female (*N*=125) | 52 (57%) | 17 (46%) | 34 (52%) | |
| Male (*N*=120) | 39 (43%) | 20 (54%) | 32 (49%) | |

**Table S2.** Demographics and summary according to age of first words. Missing data are not displayed. [1]Results of chi-square tests for categorical variables. IED: interictal epileptiform discharge, TV: truncating variant.

| Variable | Groups | | | *P* value[1] |
|---|---|---|---|---|
| **Age of walking** | **Below 18 months** | **After 18 months** | **Never achieved** | |
| ***De novo*** | | | | <0.001 |
| *De novo* (*N*=133) | 30 (32%) | 41 (84%) | 55 (100%) | |
| Inherited (*N*=89) | 65 (68%) | 8 (16%) | 0 (1%) | |
| **Exon** | | | | <0.001 |
| 1-2 (*N*=14) | 8 (7%) | 3 (5%) | 0 (0%) | |
| 3-5 (*N*=79) | 21 (18%) | 21 (38%) | 26 (40%) | |
| 6-7 (*N*=54) | 7 (6%) | 11 (20%) | 34 (51%) | |
| 8-11 (*N*=18) | 12 (10%) | 4 (7%) | 0 (0%) | |
| 12-17 (*N*=43) | 19 (16%) | 13 (23%) | 5 (6%) | |
| TV (*N*=69) | 50 (43%) | 4 (7%) | 2 (2%) | |
| **Hotspot** | | | | <0.001 |
| Non hotspot (*N*=55) | 30 (26%) | 9 (16%) | 5 (6%) | |
| Hotspot (*N*=153) | 37 (32%) | 42 (76%) | 61 (92%) | |
| TV (*N*=69) | 50 (43%) | 4 (7%) | 2 (2%) | |
| **Seizure onset age** | | | | 0.020 |
| First day (*N*=70) | 15 (13%) | 24 (44%) | 29 (45%) | |
| 2nd day – 1 month (*N*=160) | 78 (68%) | 27 (50%) | 28 (44%) | |
| After 1 month / never (*N*=40) | 22 (19%) | 3 (6%) | 7 (11%) | |
| **Seizure onset type** | | | | 0.001 |
| Sequential (*N*=123) | 78 (67%) | 15 (28%) | 17 (28%) | |
| Tonic (*N*=91) | 19 (16%) | 35 (66%) | 33 (54%) | |
| Other / no seizures (*N*=45) | 19 (16%) | 3 (6%) | 11 (18%) | |
| **EEG at seizure onset** | | | | <0.001 |
| Normal (*N*=39) | 28 (46%) | 6 (13%) | 2 (4%) | |
| Burst suppression (*N*=42) | 3 (5%) | 14 (30%) | 20 (36%) | |
| (Multi)focal IEDs with/without background slowing (*N*=95) | 30 (49%) | 26 (57%) | 31 (60%) | |
| **Frequency of seizures at onset** | | | | 0.009 |
| Multiple per day (*N*=167) | 67 (65%) | 41 (84%) | 45 (90%) | |
| Less than multiple per day (*N*=65) | 36 (35%) | 8 (16%) | 5 (10%) | |
| **Hypotonia at birth** | | | | <0.001 |
| No (*N*=156) | 100 (86%) | 16 (33%) | 9 (15%) | |
| Yes (*N*=106) | 16 (14%) | 33 (67%) | 53 (86%) | |
| **Prematurity** | | | | 0.892 |
| Present (*N*=26) | 11 (90%) | 5 (89%) | 6 (88%) | |
| Absent (*N*=200) | 95 (10%) | 39 (11%) | 42 (13%) | |
| **Sex** | | | | 0.754 |
| Female (*N*=125) | 58 (54%) | 20 (41%) | 31 (60%) | |
| Male (*N*=120) | 49 (46%) | 29 (59%) | 21 (40%) | |

**Table S3.** Demographics and summary according to age of walking. Missing data are not displayed. [1]Results of chi-square tests for categorical variables. IED: interictal epileptiform discharge, TV: truncating variant.

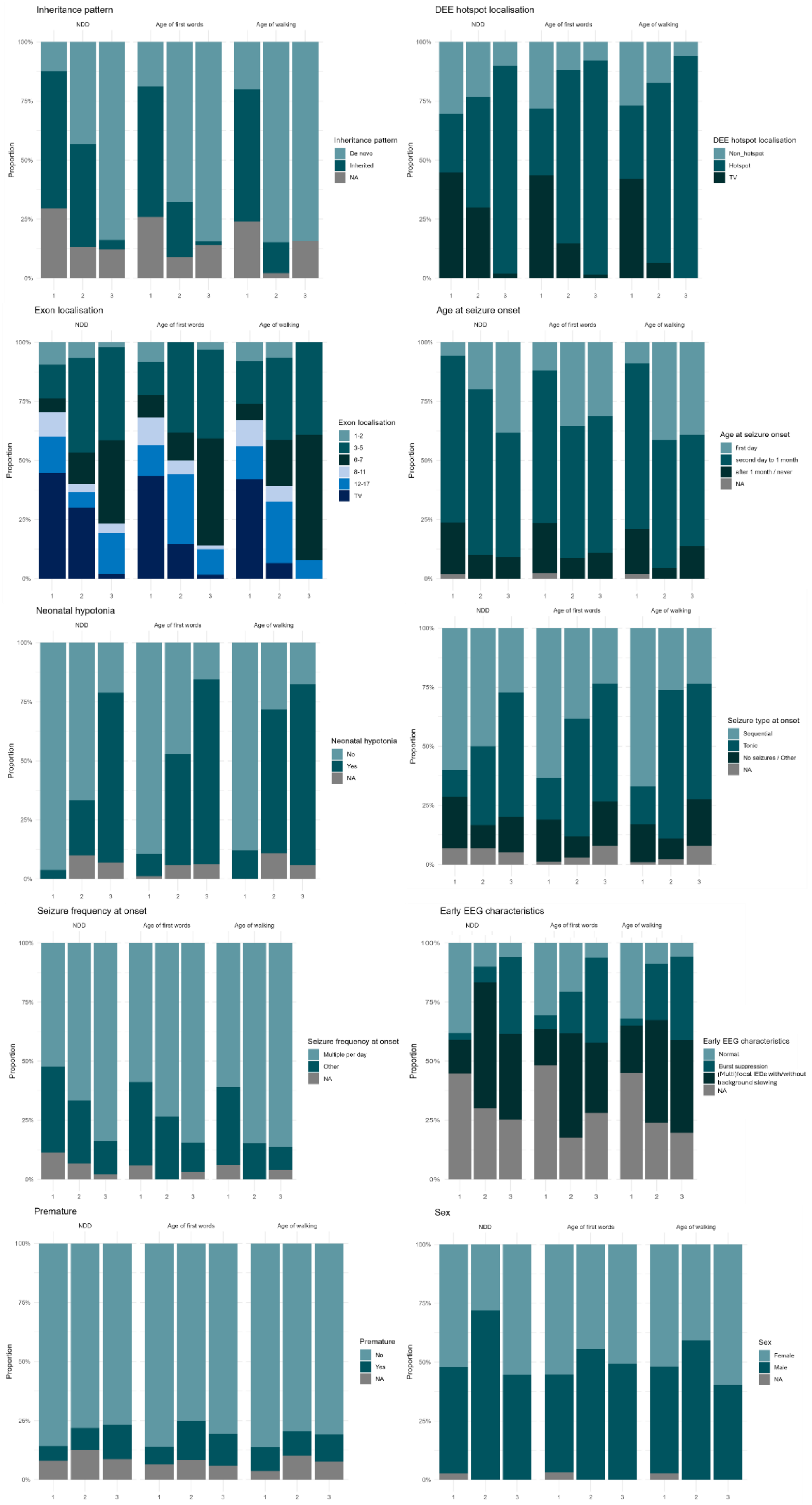
Inheritance pattern
De novo
Inherited
NA
DEE hotspot localisation
Non_hotspot
Hotspot
TV
Exon localisation
1-2
3-5
6-7
8-11
12-17
TV
Age at seizure onset
first day
second day to 1 month
after 1 month / never
NA
Neonatal hypotonia
No
Yes
NA
Seizure type at onset
Sequential
Tonic
No seizures / Other
NA
Seizure frequency at onset
Multiple per day
Other
NA
Early EEG characteristics
Normal
Burst suppression
(Multi)focal IEDs with/without background slowing
NA
Premature
No
Yes
NA
Sex
Female
Male
NA
NDD
Age of first words
Age of walking
Proportion
100%
75%
50%
25%
0%
1
2
3

**Figure S1.** Distribution of clinical and genetic predictors according to neurodevelopmental outcome categories: neurodevelopmental delay (1: normal intelligence, 2: mild ID, 3: moderate to severe ID), age of first words (1: before 16 months of age, 2: after 16 months of age, 3: never achieved talking) and age of walking (1: before 18 months of age, 2: after 18 months of age, 3: never achieved walking). TV: truncating variant, DEE: developmental and epileptic encephalopathy, IED: interictal epileptiform discharge.

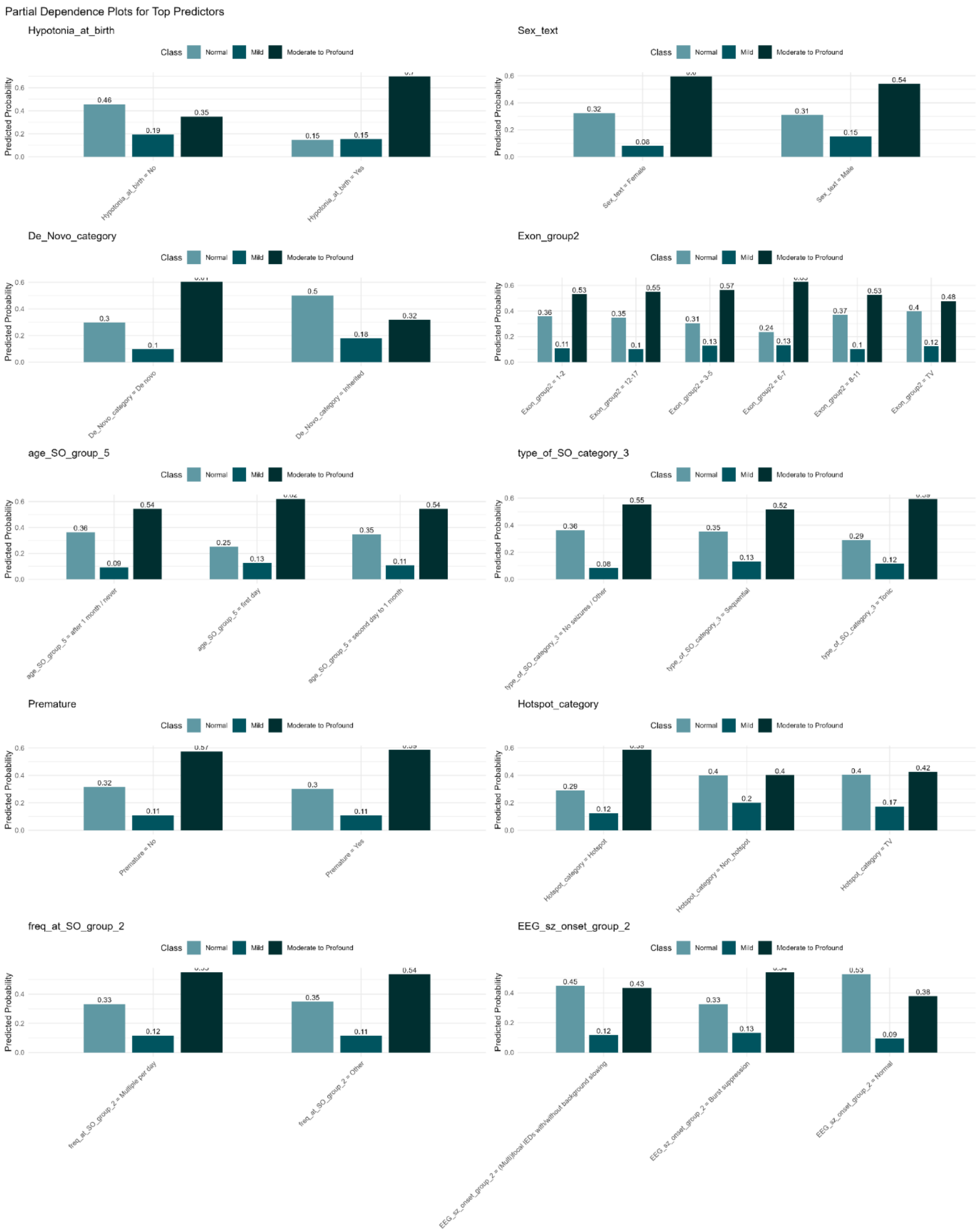


**Figure S2**. Partial dependence plots (PDPs) for the ten predictors across the random forest model predicting neurodevelopmental delay.

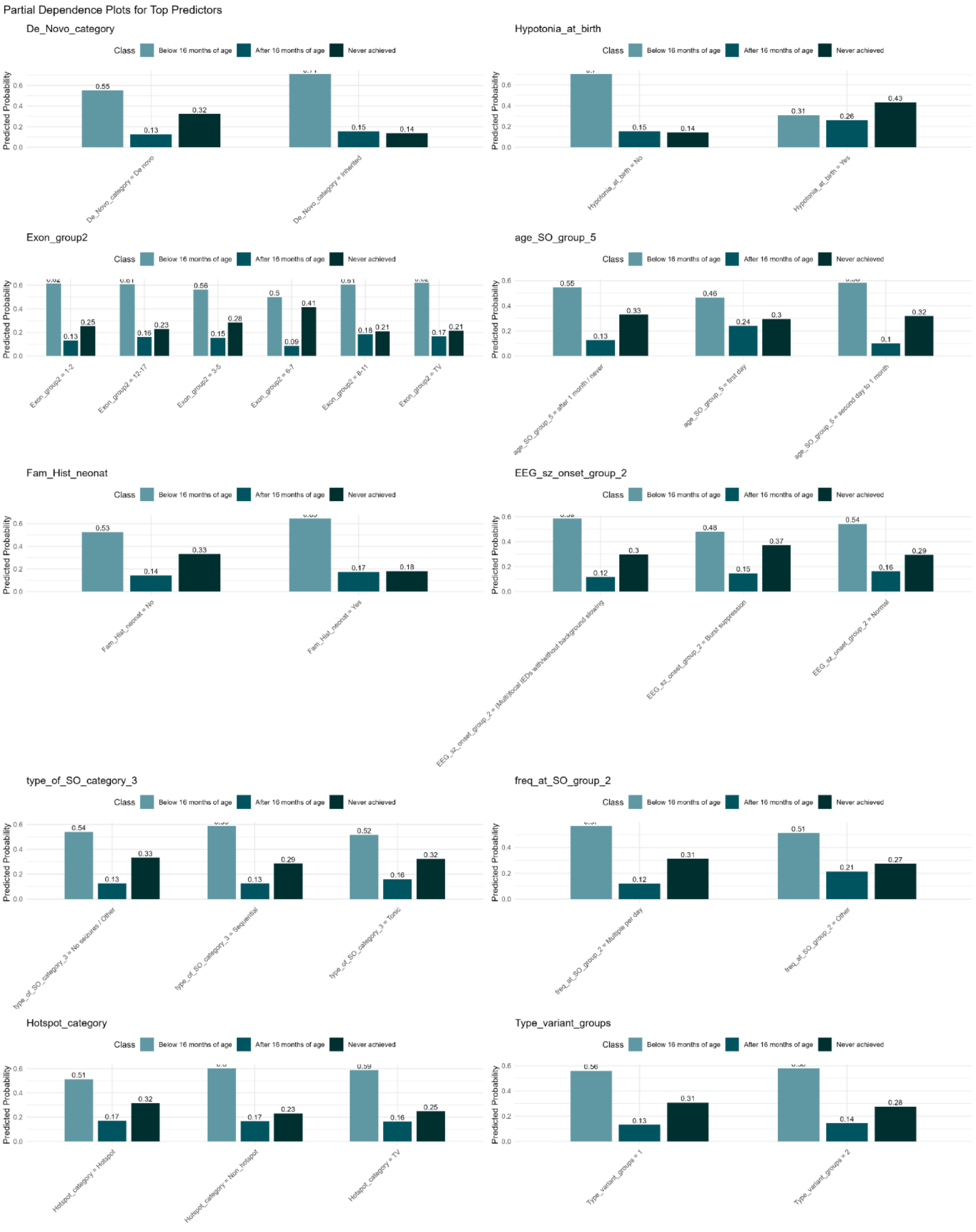


**Figure S3**. PDPs for the ten predictors across the random forest model predicting age of first words.

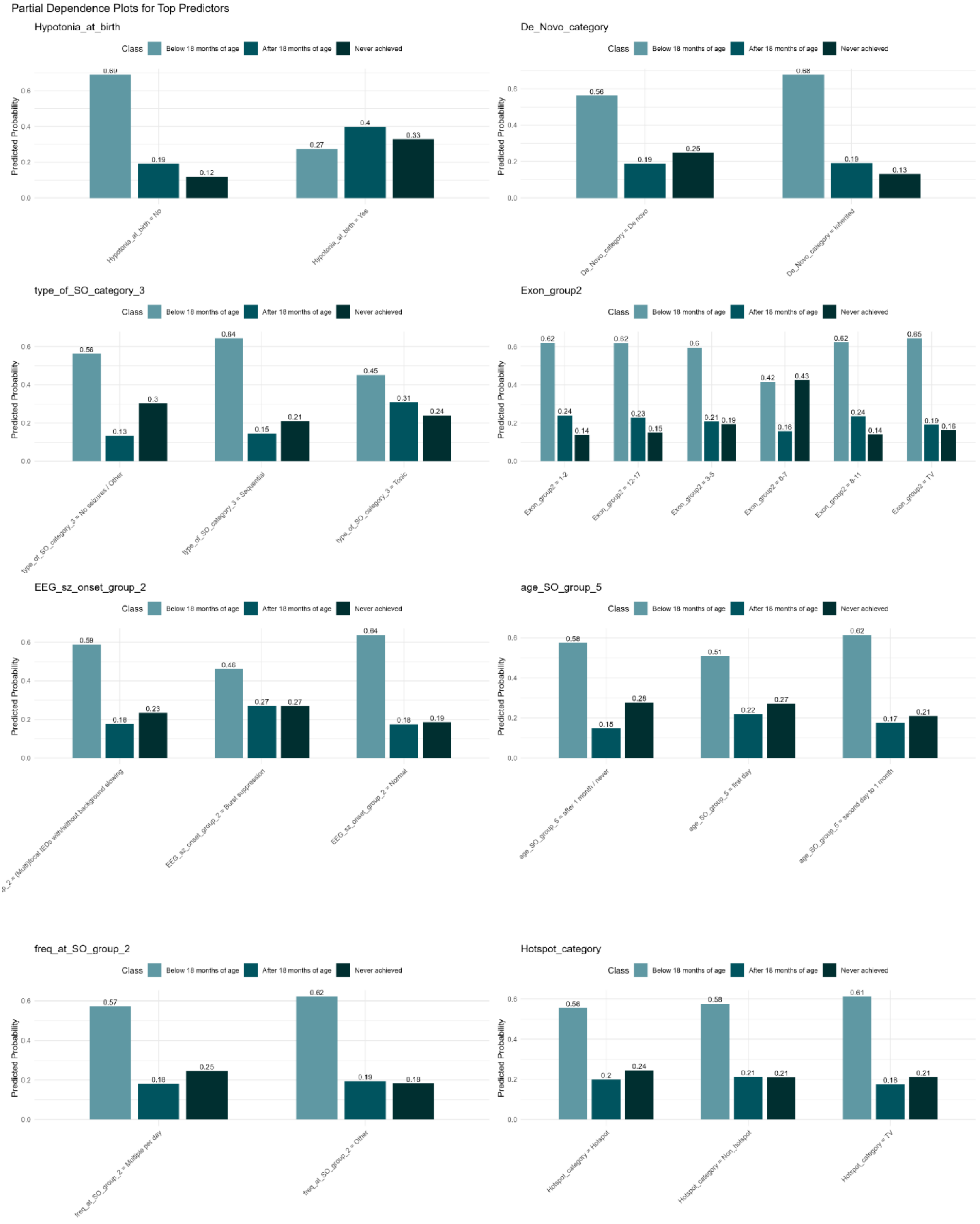


**Figure S4**. PDPs for the ten predictors across the random forest model predicting age of walking.

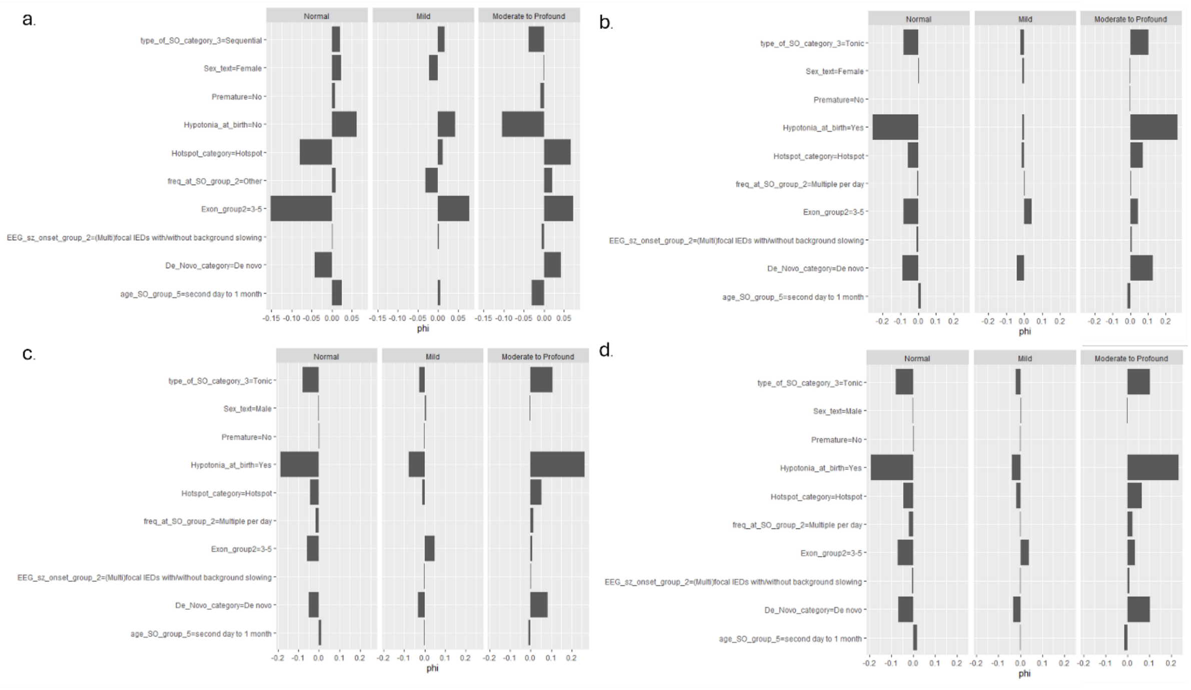


**Figure S5.** Shapley Additive exPlanations (SHAP) analysis of four patients carrying the recurrent *KCNQ2*-DEE-associated variant p.Arg213Trp and moderate to profound ID, all of whom were correctly classified by the random forest model. The outcome prediction for patient a. was lower than for patients b-d (0.43 vs. >0.90), because of factors predictive for normal intellectual outcome such as the absence of hypotonia at birth and seizure onset between two days and one month of age. However, the genetic parameters (exon group, hotspot, *de novo*). Patients b-d presented with hypotonia at birth, the major parameter for predicting more severe outcome.

**Sensitivity analysis: exclusion GoF variants, seizure onset >31 days. Neurodevelopmental delay**

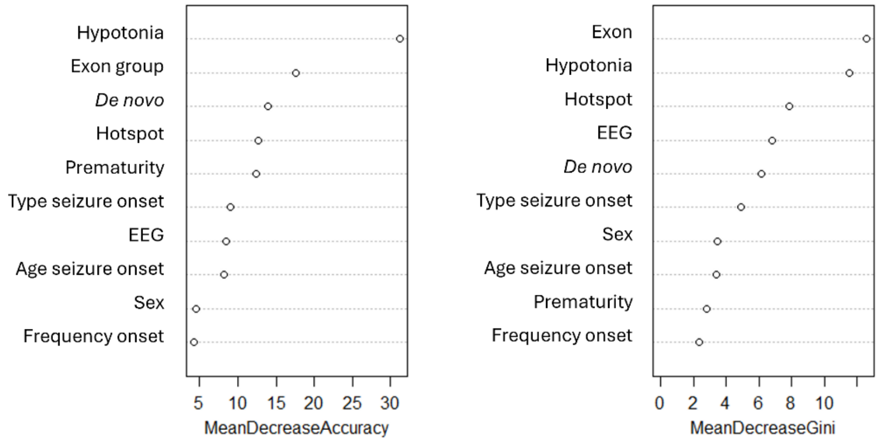


**Figure S6.** Variable importance in Random Forest sensitivity analysis model for neurodevelopmental delay, after exclusion of patients with a gain-of-function (GoF) variant, and with seizure-onset after the neonatal period (31 days), or no seizures (*n* = 184). Importance of each predictor is expressed as the mean decrease in accuracy (MDA) and mean decrease in Gini index (MDG) across all trees in the model trained on the clinically restricted cohort.

| Variable | Available, n (%) | Missing, n (%) |
|---|---|---|
| Neurodevelopmental delay | 277 (100%) | 0 (0%) |
| Age of first words | 262 (95%) | 15 (87%) |
| Age of walking | 240 (87%) | 37 (13%) |
| *De novo* | 222 (80%) | 55 (20%) |
| Exon | 277 (100%) | 0 (0%) |
| Hotspot | 277 (100%) | 0 (0%) |
| Seizure onset age | 270 (98%) | 7 (3%) |
| Seizure onset type | 259 (94%) | 18 (7%) |
| EEG at seizure onset | 176 (64%) | 1 (0%) |
| Frequency of seizures at onset | 232 (84%) | 45 (16%) |
| Hypotonia at birth | 262 (95%) | 15 (5%) |
| Prematurity | 226 (82%) | 51 (18%) |
| Sex | 245 (88%) | 32 (12%) |

**Table S4.** Missing data in the outcome and input features of the random forest models.
